# Distinct kinetics in antibody responses to 111 *Plasmodium falciparum* antigens identifies novel serological markers of recent malaria exposure

**DOI:** 10.1101/2020.12.14.20242768

**Authors:** V Yman, J Tuju, M T White, G Kamuyu, K Mwai, N Kibinge, M Asghar, C Sundling, K Sondén, L Murungi, D Kiboi, R Kimathi, T Chege, E Chepsat, P Kiyuka, L Nyamako, F H A Osier, A Färnert

## Abstract

Strengthening malaria surveillance is a key intervention needed to reduce the global disease burden. Reliable serological markers of recent malaria exposure could dramatically improve current surveillance methods by allowing for accurate estimates of infection incidence from limited data. We studied the IgG antibody response to 111 *Plasmodium falciparum* proteins in travellers followed longitudinally after a natural malaria infection in complete absence of re-exposure. We identified a novel combination of five serological markers (GAMA, MSP1, MSPDBL1 C- and N-terminal, and PfSEA1) that detect exposure within the previous 3-months with >80% sensitivity and specificity. Using mathematical modelling, we examined the antibody kinetics and determined that responses informative of recent exposure display several distinct characteristics: rapid initial boosting and decay, less inter-individual variation in response kinetics, and minimal persistence over time. These serological exposure markers can be incorporated into routine malaria surveillance to guide efforts for malaria control and elimination.

## Introduction

Reducing the global burden of malaria with the aim of achieving local or regional elimination will require sustained efforts for malaria control ^1^. This includes the implementation and the maintenance of high quality malaria surveillance systems that allow control programs to effectively allocate limited resources in their efforts to reduce disease transmission ^2,3^. Malaria surveillance has traditionally been based on estimates of the number of clinical cases reported through health systems supplemented by data on the local prevalence of infection collected through cross-sectional surveys ^4^. However, estimates of infection incidence based on passive case detection of clinical cases are heavily influenced by the quality of diagnosis and completeness of reporting and are unreliable if asymptomatic infections are common. Furthermore, the usefulness of prevalence surveys decreases substantially as transmission declines and becomes increasingly heterogeneous ^5–7^.

Antibody responses are maintained beyond the duration of actual infections and may serve as sensitive markers of past pathogen exposure that can complement traditional surveillance data. Serology has been highlighted as a useful tool for the surveillance of a wide range of infectious diseases, e.g. dengue fever, trachoma, onchocerciasis, malaria and recently COVID-19 where it is being evaluated by public health agencies worldwide ^8–11^. For malaria, serological surveillance has proven particularly useful in low transmission settings and antibody responses to a number of *Plasmodium falciparum* antigens, from both pre-erythrocytic and blood-stages, have been evaluated as exposure markers ^12–15^. In particular the responses to merozoite surface protein (MSP) 1 and apical membrane antigen 1 (AMA1) have been found to provide reliable population-level estimates of medium and long-term transmission trends ^13,16–18^. The responses to these antigens on both population and individual level, however, appear less sensitive to short-term changes in malaria transmission and reliable serological markers of recent exposure are currently lacking ^18,19^. A serological tool that provides information on the magnitude of the individual-level exposure as well as the time frame within which the individual was last exposed could improve surveillance by allowing for estimation of infection incidence from single time-point cross-sectional data ^20^. Such information could be used to monitor transmission intensity and dynamics, trigger intensified surveillance with focused malaria testing and treatment, guide targeted interventions (e.g. using long lasting insecticidal nets or other vector control measures) and subsequently evaluate their impact, or even to demonstrate the absence of transmission (reviewed in Greenhouse et al. 2018 and 2019) ^21,22^.

On the individual level, the magnitude of the malaria-specific antibody response is highly affected by both the time since last infection and the level of prior exposure ^23,24^. Although the response is generally considered to be short-lived, accumulating data further suggest that the kinetics and the longevity of the response may vary between antigens ^23,25–27^. These observations provide a rationale for attempting to identify a combination of antigens to which the antibody responses display distinct kinetics following infection (i.e. some that are short-lived and others that are more long-lived) and allow for accurate estimation of the timing of the individuals last exposure. However, an effective tool for serological surveillance would have to be limited to include only a few antigens in order to be cost-effective and feasible to implement at scale. Identifying the optimal combination of antigens will require a thorough understanding of the kinetics of each candidate antibody response. Given the scarcity of available data on antimalarial antibody kinetics, efforts should preferably start from screening a large number of candidate antigenic targets for suitability ^26,28,29^.

To date, only a few studies have attempted to identify markers for individual-level exposure, either by analysing cross-sectional data on antibody reactivity in longitudinally monitored individuals in endemic areas ^26,30–33^ or by analysing longitudinal data on antibody responses obtained from infected individuals participating in controlled human malaria infection (CHMI) trials ^34^. Helb et al. used a machine learning approach to identify candidate serological markers of recent infection by analysing cross-sectional data on antibody responses to 655 *P. falciparum* antigens collected at the end of a one-year follow-up of children monitored actively (monthly or three-monthly) and passively for parasitaemia and symptomatic infections, respectively, using microscopic examination of blood slides in an attempt to determine the timing of the last exposure prior to sampling ^26^. However, in an endemic setting this approach is notoriously difficult due to undetected exposure and a high frequency of asymptomatic carriage of low-density sub-microscopic infections ^35^. Although the timing of exposure can be carefully controlled using CHMI, participants in such trials are typically treated at microscopic or PCR patency of blood-stage infection ^36,37^ and the immune response observed may not reflect the response following a natural infection ^38^. Furthermore, CHMI studies of only primary infections ^34^ will not capture the effect that repeated parasite exposure may have on antibody profiles and kinetics ^24^. It is possible that these uncontrolled factors may have impacted which candidate serological markers have previously been suggested ^26,30,31,34^.

With the purpose of studying the acquisition and maintenance of both humoral and cell-mediated immunity to malaria, we have established a well characterised cohort of returning travellers (with different levels of prior malaria exposure) who are followed longitudinally in a malaria free country after successful treatment of a naturally acquired *P. falciparum* infection ^24,39–41^. In contrast to the design of the study by Helb et al. ^26^, samples are collected longitudinally after a known time-point of symptomatic infection. This study design offers a unique opportunity to examine the kinetics of antimalarial immune responses in complete absence of re-exposure. With this near-experimental set-up, we use a newly developed protein microarray (KILchip v1.0 ^42^) including 111 *P. falciparum* blood-stage antigens to determine the antigen-specificity and kinetics of the antibody response. We identify novel serological markers of recent malaria exposure and demonstrate how their ability to detect recent exposure depends on the underlying kinetics of each antibody response.

## Results

Sixty-five adults diagnosed with *P. falciparum* malaria at Karolinska University Hospital in Sweden were enrolled at the time of diagnosis and followed prospectively with repeated blood sampling (i.e. at enrolment, after approximately ten days, and after one, three, six, and twelve months) for up to one year in complete absence of re-exposure. Out of the 65 participants, 21 were European natives with no prior history of malaria infection who reported a limited time spent in malaria endemic areas and were considered primary infected. The remaining 44 participants (39 born in Sub-Saharan Africa) reported prior malaria episodes, and prolonged residency in malaria endemic areas, and were considered previously exposed. The two exposure groups did not differ with regards to age, sex, time from symptom onset to diagnosis, parasitaemia, or symptoms of severe malaria (Supplementary Table S1). Antibody responses to 111 *P. falciparum* blood-stage antigens were quantified in all collected sample-series using the recently developed KILchip protein microarray (KILchip v1.0). Antibody responses were largely positively correlated (Supplementary Fig. S1) and while many proteins appeared to be highly antigenic only low-level responses were observed towards others (Fig. 1). As expected, the kinetics of the antibody response was antigen-specific but on average the magnitude of the antibody response increased following the acute infection until approximately day 10 (Fig. 2a). After day 10, there was a gradual reduction in the magnitude of the response over time throughout the remainder of the follow-up period. On average, individuals with prior malaria exposure displayed a greater magnitude of the response (Fig. 2a). A similar pattern was observed for the breadth of the response (i.e. the number of antigens to which an individual is seropositive), with the peak in breadth occurring aproximately 10 days after the acute infection (primary infected: median = 17, range 7-71; previously exposed: median = 26, range 11-77) (Fig. 2b). Although none of the participants were seropositive for all antigens at any time-point, a majority of previously exposed individuals acquired and maintained a substantially greater breadth of the response at the end of follow-up (primary infected: median = 2, range 0-10; previously exposed: median 3, range 0-42) (Fig. 2b).

**Fig. 1.**
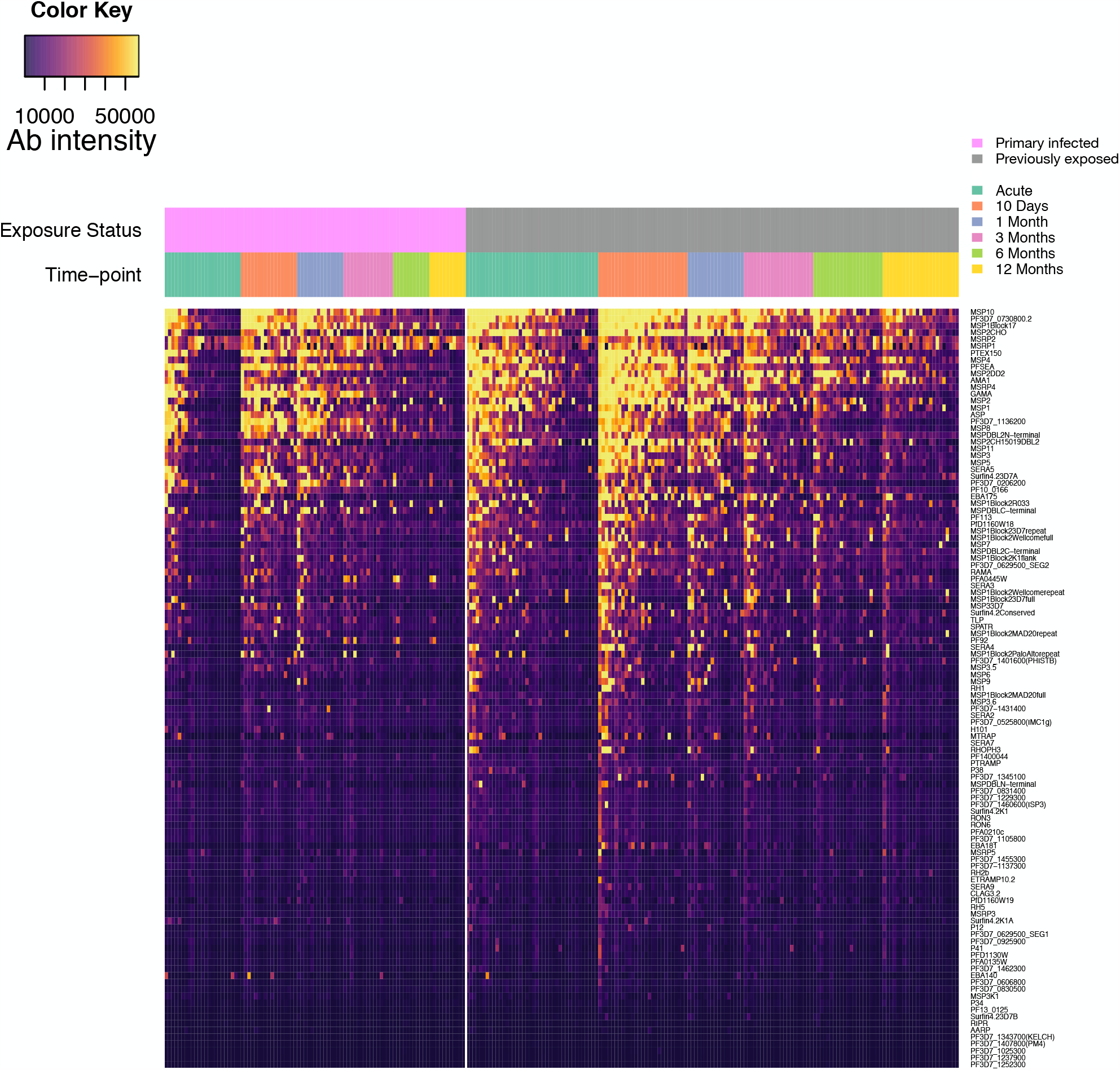
A heat map of the normalized median fluorescent intensity (MFI) of the antibody response to each of the 111 antigens included on the KILChip V1.0 Microarray. Rows correspond to individual antigens while columns correspond to individual samples. Antigens are sorted from top to bottom by decreasing average normalised MFI across all samples. Samples are sorted first by exposure status, second by sampling time-point and third by average normalised MFI across all antigens.

**Fig. 2.**
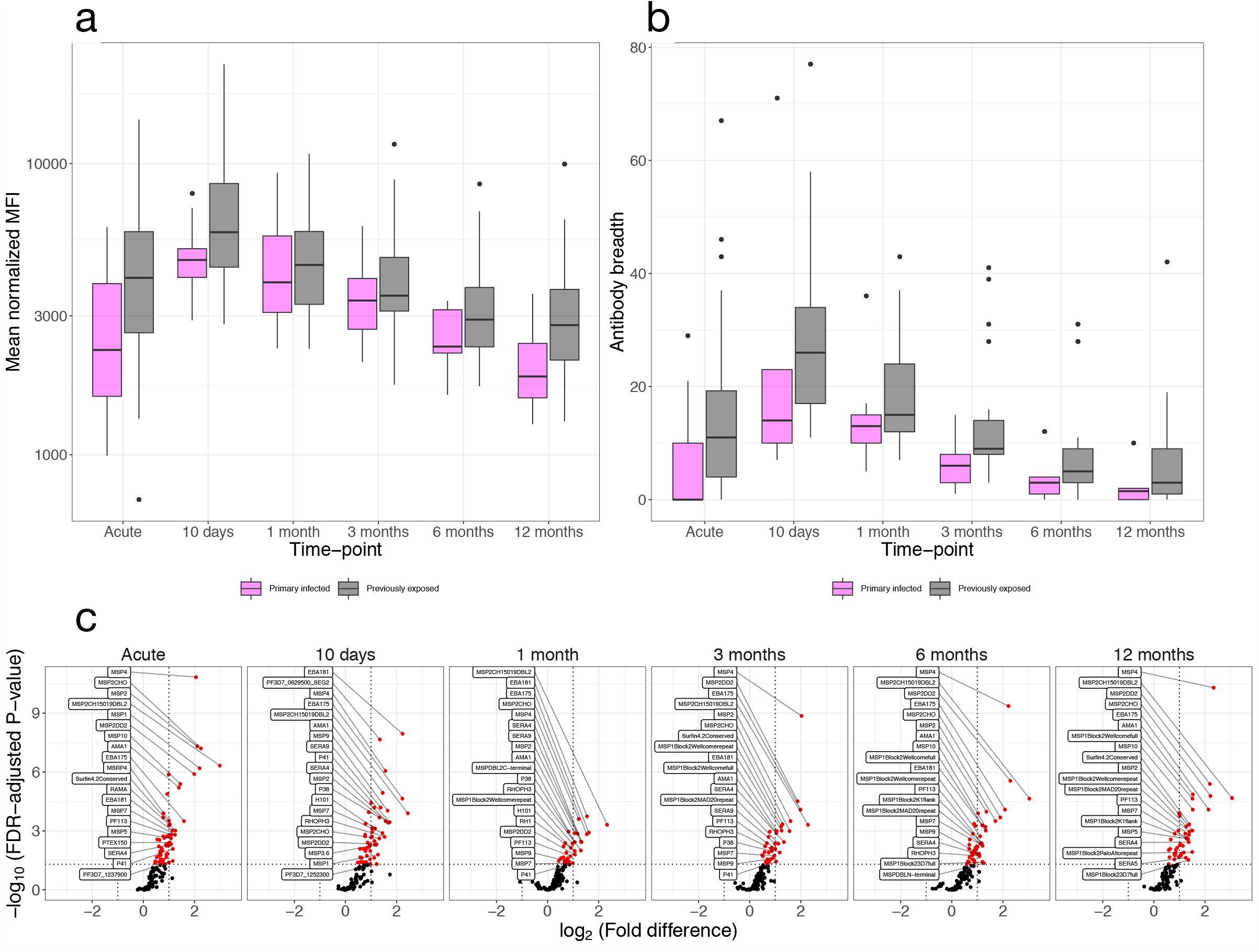
Differences in average antibody reactivity related to prior exposure. **a)** Box-plot of the overall magnitude of the antibody response to *P. falciparum* over time (averaging signal intensities over all antigens for each individual) in individuals with (grey) or without (magenta) prior malaria exposure. **b)** Box-plot of the breadth of the response over time in individuals with (grey) or without (magenta) prior malaria exposure. The breadth is expressed as the total number of antigens (out of the 111) to which the individual responds. **c)** Volcano plot of the fold-difference in geometric mean antibody reactivity between individuals with or without prior malaria exposure vs. the FDR-adjusted P-value at each of the sampling time-points. Linear mixed-effects regression models fitted to the Log-transformed antibody data were used to estimate the mean fold-difference of the response for each antigen between primary infected and previously exposed participants. P-values were FDR-adjusted for multiple comparisons using the procedure described by Benjamini and Hochberg. A log_2_ (fold difference) of greater than 0 indicates antigens to which the geometric mean response is greater among previously exposed individuals and conversely a log_2_ (fold-difference) of less than 0 indicates antigens to which the geometric mean response is greater among primary infected individuals. Antibody responses that differ significantly between exposure groups are highlighted in red and the 20 antigens for which the difference is greatest are named in the figure. Further details of this comparison are included within the supplementary information (Supplementary Table S2).

### Exposure-dependent differences in the magnitude and antigen-specificity of the response

Linear mixed-effects regression models were used to examine differences in the magnitude of the antigen-specific responses between the primary infected and the previously exposed individuals. The previously exposed individuals displayed significantly greater reactivity than the primary infected individuals toward 56 of the 111 antigens at the time of diagnosis, 54 at day 10, 32 at 1 month, 37 at three months, and 44 antigens at both 6 and 12 months of follow-up (Fig. 2c). Compared to primary infected individuals, individuals with prior exposure displayed a greater magnitude of the response to AMA1, erythrocyte-binding antigen (EBA) 175, EBA181, H101, MSP1 Block 2 (K1, MAD20, and RO33 like types) and MSP2 (full-length as well as FC27 and 3D7 type fragments) antigens, as well as to MSP4, MSP7, MSP9, merozoite surface protein duffy binding-like (MSPDBL) 2 C-terminal, Pf113, P38, RhopH3, serine repeat antigen (SERA) 3, SERA4, SERA7, and SERA9 at all time-points (Fig. 2c, Supplementary Table S2). In general, the fold-difference in antibody reactivity was greater at diagnosis and at the end of the follow-up (Fig. 2c). The observed differences, particularly at diagnosis and at the end of follow-up, indicate a more rapid induction of antibody production and better antibody maintenance upon antigenic re-stimulation suggesting the development of a memory response towards these antigens with repeated exposure. The differences in the magnitude of the response at each of the time-points are presented in full within the supplementary material (Supplementary Table S2).

### Individual antibody responses most informative of recent exposure

What is considered a recent exposure to infection may vary depending on the epidemiological setting and the purpose of a particular investigation but, in the context of *P. falciparum*, this is often defined as exposure having occurred within the past 3 to 6 months ^22,43^. For the main analysis, samples were treated as independent and a recent exposure was defined as the infection having occurred within 3 months (i.e. 90 days) of sample collection. Consequently, samples collected within 3 months of the acute infection were categorised as obtained from individuals recently exposed to infection whereas the remaining samples were not. This enabled the analysis of a balanced number of samples collected both before (52.5%) and after (47.5%) this temporal threshold within the one-year follow-up. Because a useful serological marker of recent exposure will need to accurately identify recently infected individuals regardless of their prior level of exposure, data from both exposure groups were analysed jointly. Receiver operating characteristics (ROC) analysis was applied to evaluate whether a threshold level of the antibody response towards a single antigen could be used to accurately classify if a given sample was obtained from a recently exposed individual. The analysis was performed separately for each antibody response and the performance of the classifiers was compared based on the classifier area under the ROC curve (AUC) (Fig. 3). Data on antibody levels towards several individual antigens were able to classify samples as obtained from individuals exposed within the past 3 months with comparable degrees of accuracy (Fig. 3). The best classification performance was obtained using the antibody response towards GPI-anchored micronemal antigen (GAMA) for which the AUC was 0.83 (95% CI: 0.76 – 0.90) reaching a sensitivity and specificity of 77%. Within this particular cohort this corresponded to an accuracy of 76% and a positive predictive value of 78% and a negative predictive value of 74%. Similar results were obtained using antibody responses towards *Plasmodium* translocon for exported proteins (PTEX) 150 (AUC= 0.82; 95% CI: 0.75 – 0.87), PF3D7_1136200 (AUC = 0.81; 95% CI: 0.75 – 0.88), and schizont egress antigen (*Pf*SEA-1) (AUC = 0.8; 95% CI: 0.73 – 0.87) for which the AUCs all exceeded 0.8 (Fig. 3, Supplementary Table S3). In addition, the response towards MSP8, apical sushi protein (ASP), PF3D7_0206200, MSP7-related protein (MSRP) 4, the 3D7 allelic variant of MSP3, and MSP1 Block 17 were among the top 10 most informative. However, for a majority of responses the classification performance was relatively poor (Fig. 3, Supplementary Table S3).

**Fig. 3.**
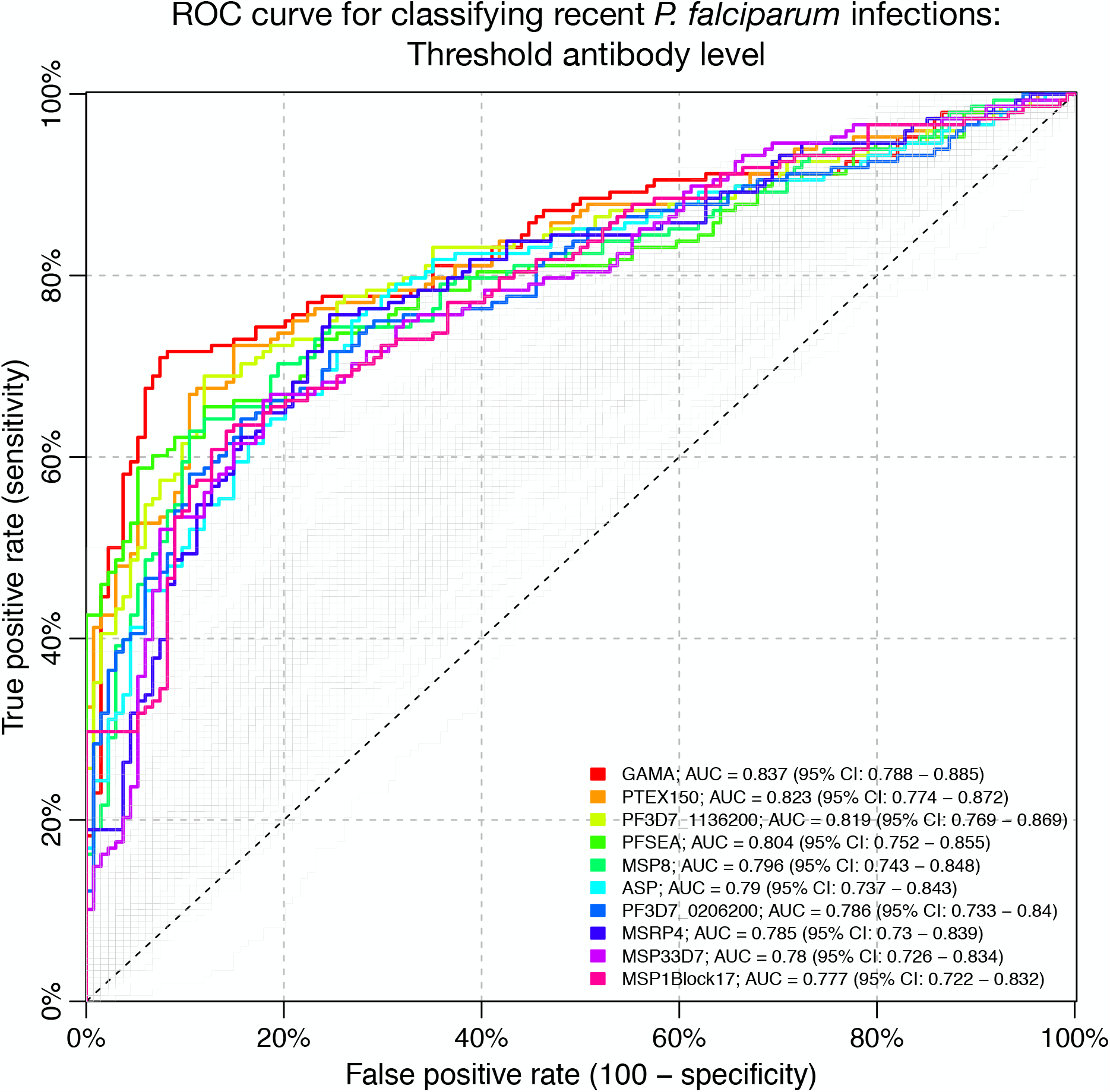
Receiver operating characteristic (ROC) curve for classifying individuals as infected within 90 days using a threshold antibody level to a single antigen. Coloured curves correspond to the top 10 antibody responses that were most accurate in detecting recent infection as determined by the classifier area under the ROC curve (AUC).

A sensitivity analysis was performed to examine whether other antibody responses would have been more informative if an alternative definition of recent exposure had been used. The analysis was therefore repeated using several definitions of a recent exposure (i.e. exposure having occurred within 1 month, 2, 3, 4, 6, and 8 months of sample collection). Although, the classifier AUCs varied depending on the definition, the same antibody responses (i.e. towards GAMA, PTEX150, MSRP4, *Pf*SEA-1, ASP, PF3D7_1136200 and MSP1 [Block 17]) were consistently identified among the top 10 responses providing most accurate identification of recent exposure (Supplementary Fig. S2).

### Combining data on multiple antibody responses to improve predictive accuracy

Combining data on antibody responses towards multiple antigens could theoretically improve the ability to accurately identify recently exposed individuals. Feature selection using a Boruta algorithm was performed to reduce the number of potential combinations to evaluate by selecting only those antibody responses contributing significant information on recent exposure when analysed together for further analysis. The algorithm, which is a wrapper algorithm based on a random forest classifier, was applied jointly to the full antibody data set. It identified 28 antibody responses contributing significant information to classification of recent exposure that were further evaluated (Fig. 4a). Similar to the results based on the threshold antibody level towards a single antigen, the Boruta algorithm identified that the greatest relative importance for classification was contributed by the response towards GAMA, *Pf*SEA1, PF3D7_1136200, PTEX150, and MSP8 (Fig. 4b). Random forest classifiers were applied to identify a panel of up to five antibody responses most informative in identifying recent exposure. The classification performance of all possible two- to five-way combinations of the 28 selected responses was exhaustively evaluated. There was a gradual increase in classifier performance, as indicated by an increase in the cross-validated AUC, with the sequential increase in panel size from two to five antibody responses. However, each increase in panel size lead to a smaller improvement in classifier performance (Supplementary Fig. S3). The antibody response to GAMA was included in all of the best combinations of two to four antibody responses (Supplementary Fig. S3). The overall best classification performance, with a cross-validated AUC of 0.89 (95% CI: 0.85 – 0.94) and reaching a sensitivity and specificity of 83%, was obtained for a panel of five antibody responses that included the response to GAMA, MSP1 (full length), both the C- and N-terminal of MSPDBL1, and *Pf*SEA1 (Fig. 4b). This corresponded to an accuracy of 83% and positive and negative predictive values of 84% and 82%, respectively. The responses to GAMA, MSP1 and the N-terminal of MSPDBL1 were included in all of the top 10 most informative panels of size five, and *Pf*SEA1 was included in 8 of the top 10 panels. The classification performance of the top 10 antibody panels was highly comparable with AUCs ranging from 0.88 (95% CI: 0.83 – 0.94) to 0.89 (95% CI: 0.85 – 0.94). The random forest classifier based on a combination of five antibody responses provided a significant improvement in classification accuracy compared to a simple classifier based on a threshold antibody level to GAMA alone (McNemar’s test: p<0.001). However, no improvement was obtained using a random forest classifier fitted jointly to data on all antibody responses (Cross-validated AUC: 0.83; 95% CI: 0.74 – 0.89). As an additional evaluation of the robustness of the results obtained using random forest classifiers, the analysis was repeated using logistic regression. The results based on logistic regression classifiers were highly comparable to those obtained using random forests and are presented within the supplementary material (Supplementary Fig. S4).

**Fig. 4.**
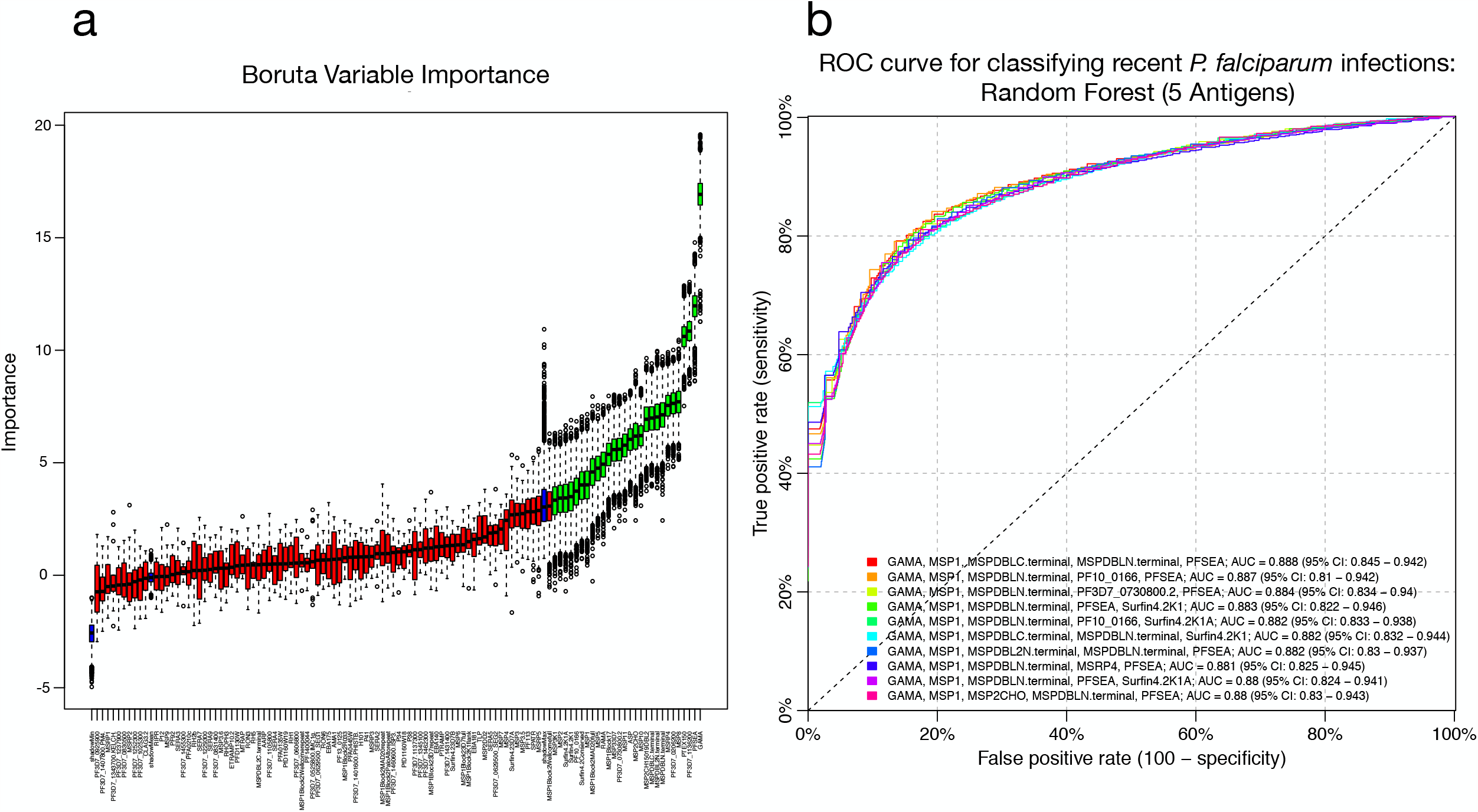
Feature selection of antibody responses and evaluation of most informative antibody combinations. **a)** Variable importance plot for classification performance determined using a Boruta feature selection algorithm as described by Kursa and Rudnicki. The Boruta algorithm was fitted jointly to data for all antibody responses. Antibody responses are ordered from left to right by their importance for classification. The importance measure is defined as the Z-score of the mean decrease accuracy (normalized permutation importance). Blue boxes correspond to the minimal, average, and maximum Z-scores of shadow features. Red boxes indicate variables not contributing significantly to accurate classification. Green boxes indicate the 28 antibody responses contributing significantly to classification that were selected for further evaluation. **b)** Cross-validated receiver operating characteristic (ROC) curves. Random forest classifiers fitted to data on antibody responses to the top 10 combinations of 5 out of the 98280 possible combinations of the 28 selected antigens as determined by the classifier area under the ROC curve (AUC). An AUC of 0.5 indicates a classifier that performs no better than random, while an AUC of 1 indicates a perfect classifier. Lines correspond the ten classifiers with the highest cross-validated AUCs.

### Identifying the antibody kinetic properties of a useful serological marker of recent exposure

Certain antibody responses (e.g. to GAMA and *Pf*SEA-1) were clearly more informative and more useful as serological markers of recent exposure than others, both independently and in combinations including multiple responses. A previously validated antibody kinetic model was applied to quantitatively describe the kinetics of each antibody response to determine if there were underlying kinetic properties shared among informative and non-informative responses. The model, which captures the inter-individual variation in boosting and decay in antibody levels following infection while estimating the average value and variance in the kinetics across the entire cohort, was fitted separately to data for each antibody response in a Bayesian framework using mixed-effects methods. The model-estimated population-level parameters and corresponding variance parameters as well as the individual level parameters are presented for all antibody responses within supplementary information (Supplementary Tables S4 and S5). An overview of the different kinetic patterns observed is presented in Fig. 5 which includes data and model fits for two representative individuals as well as the model-estimated population-averaged kinetics of the responses towards three antigens, GAMA, EBA175, and PF3D7_1252300, which were identified as highly, moderately, and minimally informative of recent exposure, respectively. The major antibody kinetic patterns observed were: i) a rapid increase and decay following infection with limited differences between individuals with and without prior exposure (Fig 5A) ii) a rapid increase and decay following infection but with substantial differences between individuals with and without prior exposure (Fig 5B) iii) a limited boosting and decay following infection with or without differences between individuals with and without prior exposure (Fig 5C).

**Fig. 5.**
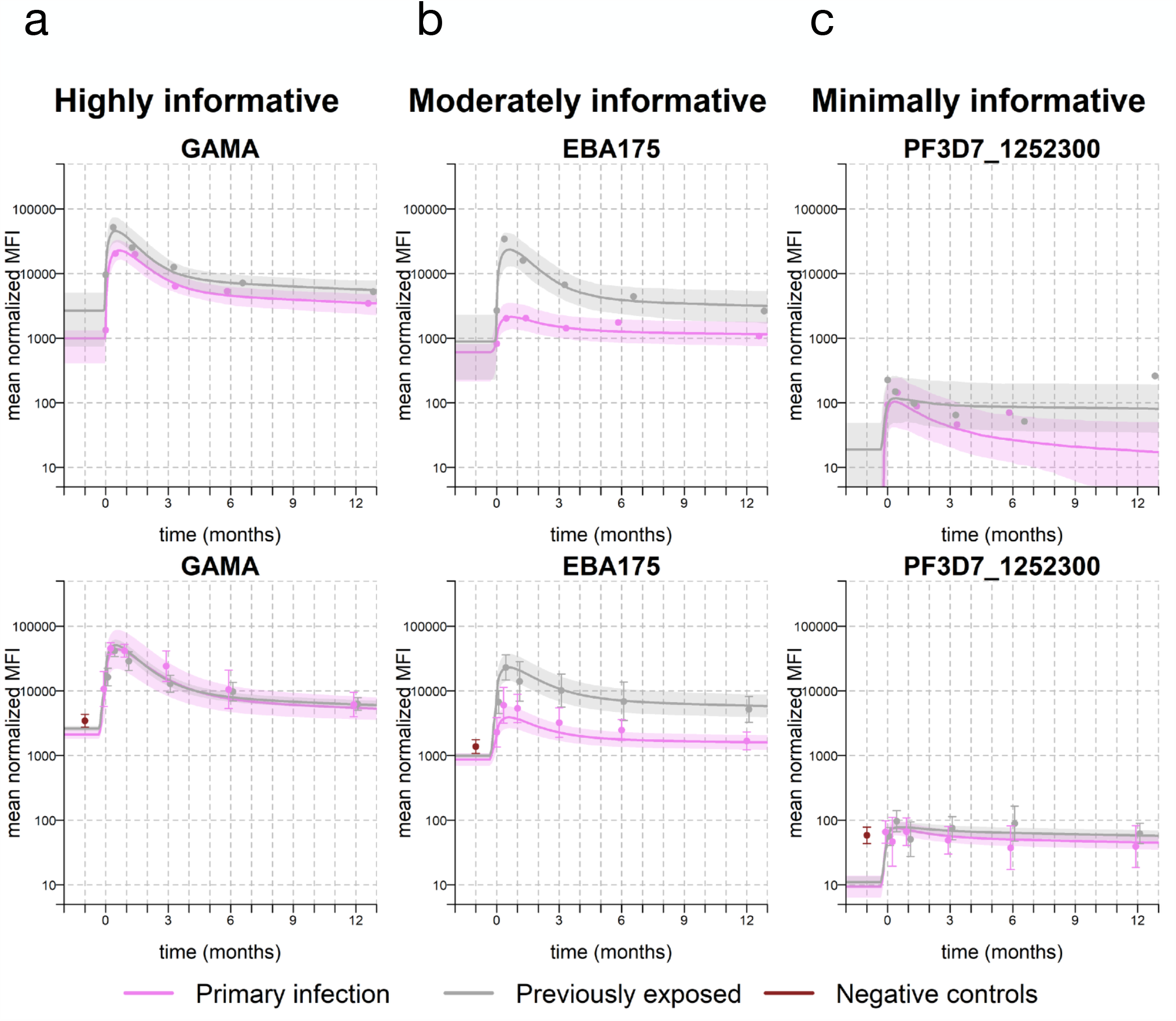
Individual- and population-level antibody kinetics for the responses to three representative antigens (GAMA, EBA175, and PF3D7_1252300) in primary infected and previously exposed individuals. The major antibody kinetic patterns observed were: **a)** a rapid increase and decay following infection with limited differences between individuals with and without prior exposure, **b)** a rapid increase and decay following infection but with substantial differences between individuals with and without prior exposure, and **c)** a limited boosting and decay following infection with or without differences between individuals with or without prior exposure. The top row displays the antibody kinetics for two representative study subjects who were either primary infected (ID: 2015006) or previously exposed (ID: 2013008). The dots denote the individual sample antibody reactivity. The solid lines denote the model predicted antibody boost and decay patterns relative to the collection of the first sample at time *t* = 0 and the shaded area the 95% credible interval of the prediction. The bottom row displays the geometric mean antibody reactivity over time in each exposure group relative to the collection of the first sample at time *t* = 0. The grey and magenta dots denote the average reactivity in previously exposed and primary infected individuals, respectively, at each sampling time point. The red dots denote average reactivity in negative control samples. The solid lines denote the model predicted mean boosting and decay in each exposure group and the shaded area the 95% confidence interval.

To be able to present a meaningful comparison of the different kinetics (i.e. the specific boosting and decay patterns) across all of the antigen-specific responses, a summary metric of the individual-level antibody kinetics for each participant and antibody response was generated by calculating the relative reduction (%) in antibody levels over the 1-year follow-up. The median relative reduction, as well as the inter-individual variation, differed substantially between antibody responses (Fig. 6). The greatest relative reductions were estimated for the highly antigenic proteins, e.g. GAMA and PF3D7_1136200, while the smallest relative reductions were estimated for poorly antigenic proteins, e.g. PF3D7_1343700.KELCH and MSRP5 (Fig. 6). All of the antibody responses that had individually been identified among the top 10 most informative in identifying recent exposure (i.e. GAMA, PTEX150, PF3D7_1136200, *Pf*SEA-1, MSP8, ASP, PF3D7_0206200, MSRP4, MSP33D7, Block 17 of MSP1) exhibited a substantial relative reduction in antibody levels during follow-up (Fig. 6). For a given antibody response, there was a close association between the estimated relative reduction in antibody levels over time and the performance (AUC) of the corresponding classifier of recent infection (Fig. 7). For the top 10 individually most informative responses, there was a limited inter-individual variation in the reduction in antibody levels over time as well as limited differences between individuals with different levels of prior malaria exposure and thus a consistent boosting and decay of the response across individuals (Fig. 6, Supplementary Fig. 5a and b).

**Fig. 6.**
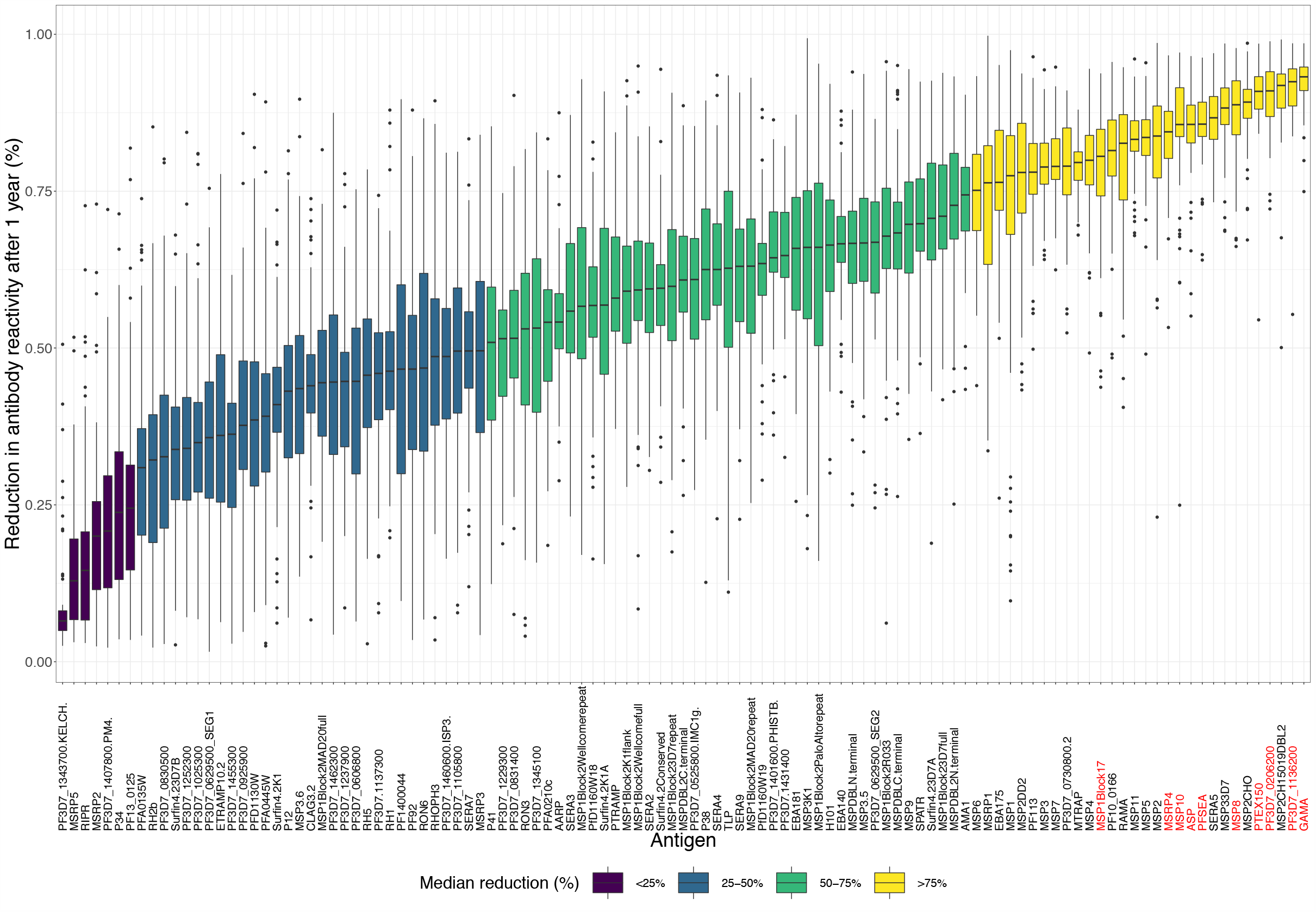
Box-plot of individual antigen-specific relative reduction (%) in antibody levels after one year of follow-up. Responses are ordered from left to right by smallest to largest relative reduction in antibody levels. The individual responses identified as top 10 most informative in detecting recent infection based on a threshold antibody level to a single antigen are highlighted in red.

**Fig. 7.**
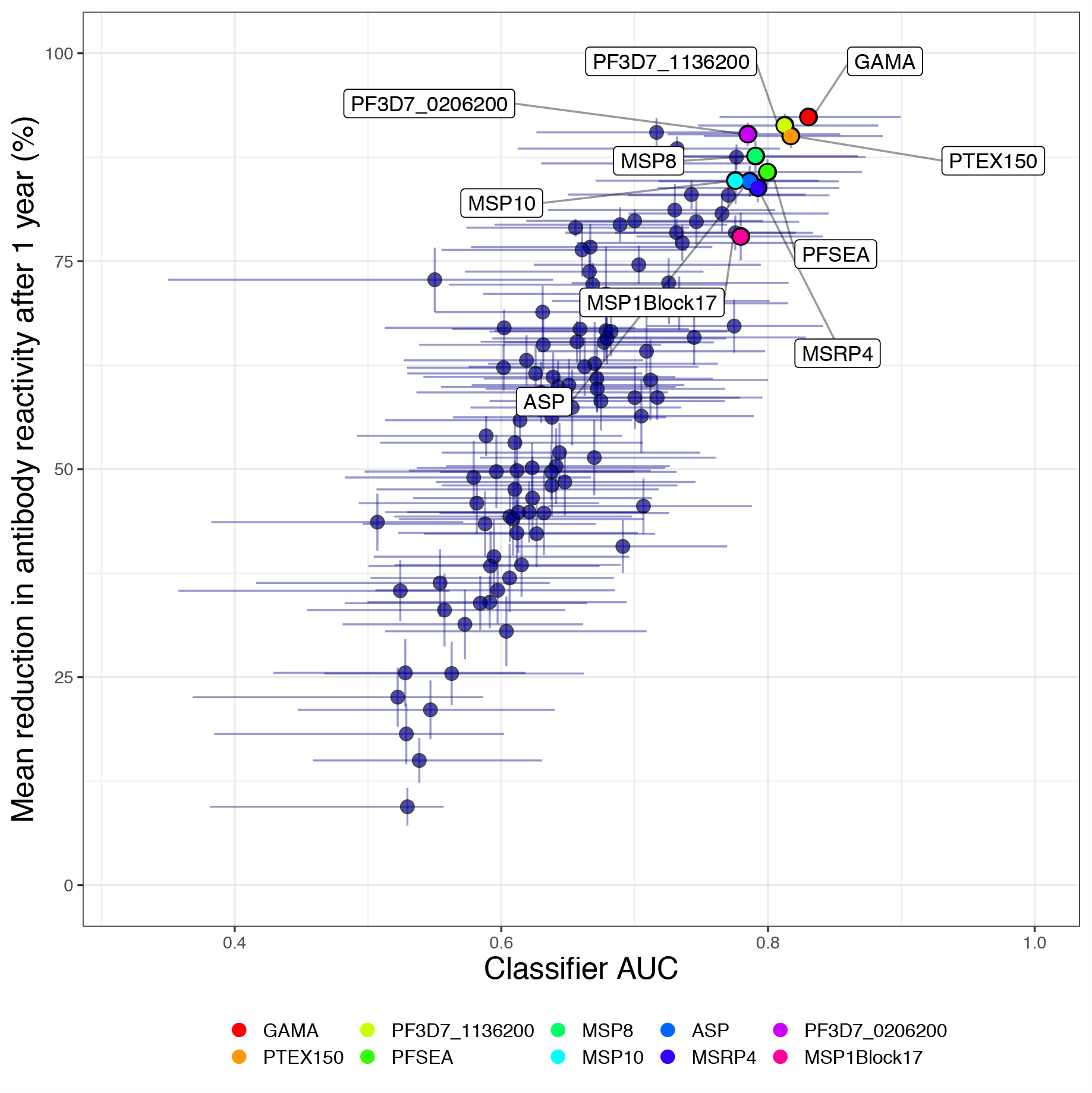
Relationship between classification accuracy and the relative reduction in antibody levels. Mean relative reduction in antibody levels after 1 year of follow-up versus classifier performance as evaluated using the classifier area under the receiver operating characteristic (ROC) curve (AUC). Colours indicate the antibody responses identified as top 10 most accurate in detecting recent infection, when using the response to a single antigen. Vertical and horizontal error bars represent the 95% confidence interval (CI) of the estimated mean relative reduction in antibody levels and classifier AUCs, respectively.

## Discussion

Novel and improved tools for malaria transmission surveillance are urgently needed to assist effective allocation of limited resources for malaria control and assure continued progress towards malaria elimination ^3^. There is a particular need for methods that can detect recent exposure to infection on the individual level which can be used to generate accurate estimates of infection incidence using limited samples and data ^20–22^. Here, we screened plasma samples from 65 travellers followed prospectively for up to one year after a naturally acquired *P. falciparum* infection for IgG antibody responses towards 111 blood-stage antigens. Using a data driven approach, we identified novel candidate serological exposure markers individually informative of recent exposure and demonstrate that combining data on five responses allow for accurate detection of recent exposure to *P. falciparum* within the prior 3-month period. Based on a modelling approach, we then quantitatively examined the kinetics of each individual antibody response and were able to characterise the kinetic properties that make a particular antibody response useful as a serological marker of recent *P. falciparum* exposure.

When examining each of the 111 antibody responses individually, we found that the level of the response to several antigens, in particular GAMA, PTEX150, PF3D7_1136200, and *Pf*SEA1, were informative and could be used to identify a recent exposure with comparable accuracy (Classifier AUCs all exceeding 0.8). The response to GAMA was most informative and it was possible to identify a threshold antibody level such that recently exposed individuals could be identified with a sensitivity and specificity of 77%. The required sensitivity and specificity of a particular surveillance system, and the optimal trade-off between them, should be dictated by the objective of the system, the activity the system is supposed to trigger, the availability of resources and cost of possible interventions ^22,44,45^. The level of accuracy in detection of recent exposure achievable using a single antibody response could be acceptable for effective serosurveillance of population-level transmission trends where e.g. a lower sensitivity can be acceptable given that it remains constant over time ^46,47^.

We demonstrated that the ability to accurately detect recent exposure could be significantly improved if data on up to five antibody responses were analysed simultaneously using a random forest algorithm. We found that the best performance was obtained based on a panel of five antibody responses (AUC = 0.89), reaching a sensitivity and specificity of 83%. There was no single best antibody combination, instead many panels composed of five antibody responses provided comparable results. All of the top 10 panels included responses that had individually been identified as highly informative (e.g. to GAMA and *Pf*SEA-1), suggesting that proteins that can identify recent infections when used individually also do well in combinations. Interestingly, however, they also included responses that were individually not among the more informative (i.e. to MSP1 and either one or both of the N- and C-terminal of MSPDBL1) suggesting that these responses contribute additional information when used in combination with individually informative responses.

The antibody responses to most of the proteins that we identified as informative of recent exposure have to date not been extensively studied. Among our top 28 candidates, which were informative either individually or in combination, only the responses to *Pf*SEA-1, PTEX150, MSP1 (19 kDa fragment and full length), MSP2 and MSP10 have to our knowledge previously been suggested as markers of recent or concurrent infection ^26,27,30,32,34,48^. The response to MSP4 and SERA4 have recently been suggested as markers of recent exposure based on data from primary infections in CHMI trials ^34,49^. However, in our study we did not find the response to MSP4 or SERA4 informative and throughout the follow-up we observed substantially greater levels of the response to these antigens in previously exposed compared to primary infected individuals. We observed similar patterns for AMA1, EBA175, EBA181, H101, MSP1 (Block 2) and MSP2 antigens, as well as to MSP7, MSP9, MSPDBL2 C-terminal, Pf113, P38, RhopH3, SERA3, SERA7, and SERA9 and believe that the prior exposure-related differences in the response to all of these antigens suggest they may also (to varying extents) reflect cumulative malaria exposure. This is corroborated by other studies where this has been extensively demonstrated for AMA1, MSP1 (Block 17) and MSP2 ^13,18,24,32,48,50,51^, and to some extent also reported for EBA181, MSP1 (Block 2), MSP4, MSP7, MSP10, and SERA4, to which the magnitude of the response increases significantly with age in populations living in areas with on-going malaria transmission ^26,30,31,51–53^. Furthermore, the differences in the antigen-specificity and breadth of the response in primary versus re-infection contribute interesting information regarding antibody acquisition and maintenance that may provide insights regarding the development of malarial immunity. However, investigating the immunological determinants of the observed differences was not among the objectives of the present study and will therefore be explored elsewhere.

It has been suggested that what determines the usefulness of any particular response as a marker of recent exposure is not just the average of its boosting or decay following infection but also the variation in these qualities across individuals ^22^. When studied individually, several antibody responses (e.g. to GAMA, PTEX150, PF3D7_1136200, *Pf*SEA-1, and MSP8) were consistently identified as the most informative in detecting recent exposure, suggesting they may share common properties with regards to their kinetics. Because of the longitudinal design of the study, we were able to examine the kinetics of each antibody response in detail using a previously validated mathematical model ^23,24^. This allowed us to quantitatively characterise both the antibody boosting and decay, its inter-individual variation as well as its dependency on prior malaria exposure and to identify three key aspects that make a particular antibody response a useful serological marker of recent exposure: i) a rapid boosting and decay in antibody levels following clearance of infection ii) limited inter-individual variation in the kinetics (boosting and decay) of the response and therefore predictable kinetics iii) minimal impact on the kinetics due to prior exposure and a limited formation of an antibody memory response. We could also show that antibody responses that were not informative of recent exposure did not exhibit this behaviour and thereby explicitly demonstrate how the ability to identify recent exposure using serology is based on an understanding of the underlying antibody kinetics.

In the present study we have intentionally focused solely on serology with the aim of identifying a panel of antibody responses that can accurately detect recent *P. falciparum* exposure without the use of additional data. In a surveillance context, data obtained from these serological markers of recent exposure could easily be combined with parasitological data as has been previously demonstrated for sero-surveillance of *P. vivax*, where Longley and White et al. identified candidate serological exposure markers that can be used to detect recent exposure infection ^33^, as well as for infectious disease surveillance in animal populations ^20^. Furthermore, if a serological assay is to be implemented at scale for field based surveillance it could preferably be developed into a multicomponent serological lateral flow immune-chromatographic point-of-care test. In such a setup, serological detection could be effectively integrated with parasite detection e.g. using ultrasensitive antigen detection ^32,54,55.^

The unique longitudinal design of this study, in which the exact time-point of natural exposure is known and where the absence of re-exposure during follow-up can be guaranteed, avoids misclassification of true exposure status thereby limiting bias and providing a unique opportunity to identify markers of recent exposure. Furthermore, including individuals who are both primary infected and previously exposed allowed us to ascertain that our candidate serological markers were able to perform equally well independently of the individuals prior level of exposure. This in turn suggests that identified candidate responses could be suitable for exposure monitoring through a wide range of transmission settings ^22,56^. We firmly believe that further evaluation and validation of these candidate serological markers of recent exposure is warranted and aim to pursue this using a range of immunoassay platforms and by performing additional validation using samples collected from longitudinally followed and closely monitored populations living in malaria endemic areas.

In summary, we identify novel candidate serological markers of recent exposure that, when quantified individually or in combination in a single plasma sample, provide information on when the donor was last exposed to *P. falciparum* infection. Using both a data driven and a modelling approach, we demonstrate that a recent exposure is not necessarily identified by a complex antibody signature that requires sophisticated algorithms for detection but rather by a thorough understanding of the kinetics of the antibody response to limited number of antigens. We show that the antibody responses towards highly antigenic proteins that, regardless of the number of prior infections, demonstrate predictable boosting and decay following a new infection are sufficient to detect whether a given individual has been exposed within a defined period of time. These novel serological markers generate information that can be used for malaria control purposes to understand when and where to intensify surveillance, perform targeted testing and treatment, and/or deploy vector control measures, and thereby effectively improve efforts to limit transmission and accelerate progress towards malaria elimination.

## Methods

### Study population

Study participants were recruited at Karolinska University Hospital in Stockholm, Sweden. Adults with *P. falciparum* malaria were enrolled at the time of diagnosis and followed prospectively for up to one year with repeated blood sampling. Venous blood samples were collected at the time of enrolment (i.e. at diagnosis) and follow-up samples were collected approximately ten days, and one, three, six, and twelve months after the first sample. In total, 242 samples were collected from 65 participants. Data on country of birth, previous countries of residence, travel history, use of antimalarial prophylaxis, previous malaria episodes and co-morbidities was collected using a questionnaire administered to each study participant upon enrolment as well as at the end of the follow-up period. Additional clinical data were extracted from hospital records ^24^.

### Ethics statement

The study was approved by the Ethical Review Board in Stockholm, Sweden (Dnr 2006/893-31/4 and 2013/550-32/4, 2018/2354-32, 2019-03436) and written informed consent was obtained from all study participants.

### Protein microarray (KILchip v1.0)

The KILchip v1.0 protein microarray was used for simultaneous quantification of IgG antibody responses to 111 *P. falciparum* antigens ^42^. The microarray includes 82 full-length proteins (or for multi-membrane proteins, the largest predicted extracellular loop) and 29 protein fragments from 8 unique proteins (i.e. MSP1, MSP2, MSP3, MSPDBL1, MSPDBL2, *Pf*SEA-1, PF3D7_06293500 and SURFIN 4.2). A majority of proteins were produced using a mammalian expression system, while a minority were produced in *Escherichia coli*. Further details regarding the microarray method are described elsewhere ^42^. Plasma samples were analysed in 1:400 dilution and bound antibody was detected using AlexaFluor^647^ conjugated donkey anti-human-IgG. The processed microarray slides were read at 635 nm using a GenePix® 4000B scanner (Molecular Devices) and results obtained using the GenePix® Pro 7 software (Molecular Devices). Positive and negative controls consisting of pooled plasma from malaria exposed Kenyan adults and serum samples from malaria unexposed northern European donors, respectively, were run on each slide. A 3-fold serially diluted standard calibrator consisting of purified IgG from highly malaria exposed Kenyan donors was assayed once within each batch.

### Data analysis

#### Data acquisition, cleaning and normalisation

R (R: A language and environment for statistical computing, v3.4.4 and v3.6.1) was used for data processing, normalization, and analyses. The median fluorescent intensities (MFI) of the local spot background surrounding each spot was subtracted from the MFI of each antigen spot. The mean MFIs of replicate spots were log-transformed to yield an approximate Gaussian distribution of signal intensities. To account for technical slide-to-slide and batch-to-batch variation a two-step normalisation process was applied according to previously described procedures ^57,58^. First, to account for within batch slide-to-slide effects, a Robust Linear Model (RLM) was fitted to the log-transformed data from the positive control samples assayed on each slide. This was done separately for data from each batch ^57^. After obtaining the best-fit parameters for the slide effect the estimated coefficients for each slide was subtracted from all spots within each slide. Following this within-batch RLM normalisation, a second between-batch RLM normalisation was performed using data for the serially diluted standard calibrator. Data for all target antigens that did not demonstrate optical saturation or no signal was used for normalisation in both steps. A threshold of seropositivity was defined as the mean reactivity + 3SD of the 42 negative controls. The breadth of the response within each tested sample was defined as the number of antigens for which the reactivity exceeded the seropositivity threshold.

#### Exposure dependent differences in geometric mean antibody responses

Linear mixed effects regression models were used to identify antigens to which responses were significantly different between primary infected and previously exposed individuals at each sampling time-point. The models were fitted separately to the log-transformed normalised MFI data for each antigen. To account for the false discovery rate (FDR) due to testing such a large number of hypotheses all p-values were FDR-adjusted according to the procedures described by Benjamini and Hochberg ^59^. FDR adjusted p-values of <0.05 were considered significant.

#### Identifying antibody responses informative of recent exposure to infection using binary classification based on single antibody responses

For the purpose of the main analysis a recent exposure was defined as the infection having occurred within 3 months (i.e. 90 days) of sample collection. All samples were categorised as obtained from either a “recently infected” or “not recently infected” individual depending on whether or not they were collected within this specified time frame. To evaluate if the antibody response to any single *P. falciparum* antigen was informative of recent exposure, binary classification using a threshold antibody level was applied to the data for each of the 111 antigens individually using ROC analysis. The AUC was used to compare the classification performance of the individual antibody responses and confidence intervals for the AUCs were estimated using DeLong’s algorithm ^60^. Alternate definitions of recent exposure were also evaluated as part of a sensitivity analysis.

#### Feature selection using a Boruta algorithm

Combining data on multiple antibody responses could theoretically improve the ability to accurately identify recent exposure, however, there are 2^111^ potential unique combinations of antibody responses to 111 antigens and to evaluate them all was not feasible ^61^. To reduce the number of tentative antibody response combinations to evaluate, feature selection was performed using a Boruta algorithm as previously described ^62^. The Boruta algorithm is a wrapper method built around a random forest classifier that performs a top-down search for relevant features, while progressively eliminating irrelevant features, by comparing the importance of original features with the importance achievable at random (estimated using permuted copies of the original features). The algorithm was fitted jointly to antibody data to all 111 antigens.

#### Identifying combinations of antibody responses informative of recent exposure to infection using random forest classification

Following feature selection, random forest classifiers were fitted exhaustively to all possible two-to five-way combinations of the down-selected antibody responses in order to evaluate whether a combination of responses could improve performance of classification of recent infection. Classifier performance was determined by the cross-validated AUC. Cross-validation was performed for each classifier using repeated random sub-sampling by iteratively and randomly splitting the data set into a training set (2/3) and a test set (1/3) ^63^. For each split the model was fitted to the training set and the predictive accuracy assessed using the test set. The results from 500 iterations were averaged to obtain a cross-validated estimate of the classifier performance and the 0.025 and 0.975 quantiles of the AUC across iterations were extracted to obtain a 95% confidence interval of the cross-validated AUC.

#### Modelling antibody kinetics

A previously validated mathematical model was used to estimate the antigen-specific antibody kinetics. The model captures the boosting and bi-phasic decay in antibody levels following infection and quantifies their inter-individual variation, while simultaneously accounting for differences in prior malaria exposure ^23,24^. Briefly the model assumes that the infection causes antibody levels to rise *τ*_0_ days before the individual presents to the hospital (where *τ*_0_ is a parameter estimated for each individual) and that *A*(*t*) is the antibody level at time *t* > *τ*_0_ and is given by the following equation (1):

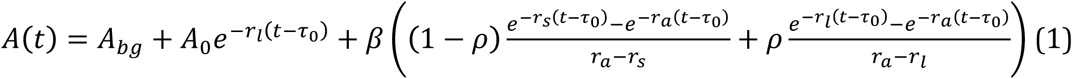

where *r*_*a*_ is the rate of decay of IgG molecules; *r*_*s*_ and *r*_*l*_ are the rates of decay of short- and long-lived antibody secreting cells (ASCs), respectively; *β* is the boost in ASCs following infection at time *τ*_0_; and *ρ* is the proportion of ASCs that are long-lived. *A*_0_ is the pre-existing levels of antibodies. For primary infected individuals, *A*_0_ = 0. *A*_*bg*_ is the background level of antibody reactivity. The models were fitted separately for each antibody response in a Bayesian framework, and mixed-effects methods were used to capture the natural variation in antibody kinetics between individuals while estimating the average value and variance of the parameters across the entire cohort. Additionally, the antibody kinetic model accounts for sample reactivity exceeding the upper limit of detection of the microarray assay. The rate of decay in antibody reactivity was expressed as the relative reduction (%) after 1 year, starting from the peak of the response ^64^.

## Data Availability

The datasets used and analysed during the current study are available from the corresponding author on reasonable request.

## Author contributions

AF, FHAO and VY planned and designed the study. VY organised the enrolment and follow-up of study participants and processed the samples together with KS, MA, and CS. GK, JT and FHAO, designed and developed the protein microarray with assistance from LM, DK, RY, EC and PK. JT, RK, and TC performed the microarray experiments. KM and NK developed the data acquisition pipeline. VY and MTW performed the data analysis, and MTW developed and fitted the antibody kinetic models. VY wrote the first draft of the manuscript. All authors contributed to critically revising the manuscript and have approved the final version.

## Acknowledgements

We are grateful to all subjects for their participation in the study. We thank Ingrid Andrén, Irene Nordling and fellow nurses at the Karolinska University Hospital, Department of Infectious Diseases outpatient ward for assistance with coordinating follow-up visits and sampling the study participants. We are grateful to Christine Stenström and colleagues at the Karolinska University Hospital, Department of Microbiology, as well as the attending physicians at the Department of Infectious Diseases, for notifying us regarding the admission of patients diagnosed with *P. falciparum* malaria.

## Supplementary Information

**File S1: Supplementary Information**. Supplementary Table S1 and Supplementary Figs. S1-S5.

**File S2: Supplementary Table S2**. Results from linear mixed effects models examining the difference in antibody levels between primary infected and previously exposed individuals. All antigens and time-points for which antibody levels were significantly different between the two exposure groups are presented within the table.

**File S3: Supplementary Table S3**. Results from ROC analysis of identifying recent exposure (within 3 months) based on a threshold antibody level for all individual antigens.

**File S4: Supplementary Table S4**. The antibody kinetic model estimated population-level parameters and corresponding variance parameters.

**File S5: Supplementary Table S5**. The antibody kinetic model estimated individual-level parameters.

## References

1. Rabinovich, R. N. et al. malERA: An updated research agenda for malaria elimination and eradication. PLoS Medicine 14, e1002456 (2017).

2. The malERA Consultative Group on Monitoring Evaluation and Surveillance. A research agenda for malaria eradication: monitoring, evaluation, and surveillance. PLoS Med. 8, e1000400 (2011).

3. World Health Organzation. Global technical strategy for malaria 2016-2030. World Health Organisation. (2015).

4. Tusting, L. S., Bousema, T., Smith, D. L. & Drakeley, C. Measuring changes in Plasmodium falciparum transmission: precision, accuracy and costs of metrics. Adv. Parasitol. 84, 151–208 (2014).

5. Hay Simon, M. E. Measuring malaria endemicity from intense to interrupted transmission. Lancet Infect. Dis. 8, 369–78 (2008).

6. Stresman, G., Cameron, A. & Drakeley, C. Freedom from Infection: Confirming Interruption of Malaria Transmission. Trends in Parasitology 33, 345–352 (2017).

7. Cooper, L. et al. Pareto rules for malaria super-spreaders and super-spreading. Nat. Commun. 10, 1–9 (2019).

8. Reiner, R. C. et al. Time-varying, serotype-specific force of infection of dengue virus. Proc. Natl. Acad. Sci. U. S. A. 111, E2694–2702 (2014).

9. Bretscher, M. T. et al. Measurement of Plasmodium falciparum transmission intensity using serological cohort data from Indonesian schoolchildren. Malar. J. 12, 21 (2013).

10. Martin, D. L. et al. Serology for trachoma surveillance after cessation of mass drug administration. PLoS Negl. Trop. Dis. 9, e0003555 (2015).

11. Golden, A. et al. Analysis of age-dependent trends in Ov16 IgG4 seroprevalence to onchocerciasis. Parasit. Vectors 9, 338 (2016).

12. Kusi, K. A. et al. Anti-sporozoite antibodies as alternative markers for malaria transmission intensity estimation. Malar. J. 13, 103 (2014).

13. Drakeley, C. J. et al. Estimating medium- and long-term trends in malaria transmission by using serological markers of malaria exposure. Proc. Natl. Acad. Sci. U. S. A. 102, 5108–5113 (2005).

14. Cook, J. et al. Using serological measures to monitor changes in malaria transmission in Vanuatu. Malar. J. 9, 169 (2010).

15. Ondigo, B. N. et al. Estimation of Recent and Long-Term Malaria Transmission in a Population by Antibody Testing to Multiple Plasmodium falciparum Antigens. J. Infect. Dis. 210, 1123–1132 (2014).

16. Cavanagh, D. R. et al. A longitudinal study of type-specific antibody responses to Plasmodium falciparum merozoite surface protein-1 in an area of unstable malaria in Sudan. J. Immunol. 161, 347–59 (1998).

17. Polley, S. D. et al. Human antibodies to recombinant protein constructs of Plasmodium falciparum Apical Membrane Antigen 1 (AMA1) and their associations with protection from malaria. Vaccine 23, 718–728 (2004).

18. Yman, V. et al. Antibody acquisition models: A new tool for serological surveillance of malaria transmission intensity. Sci. Rep. 6, 19472 (2016).

19. Wong, J. et al. Serological markers for monitoring historical changes in malaria transmission intensity in a highly endemic region of Western Kenya, 1994-2009. Malar. J. 13, 451 (2014).

20. Borremans, B., Hens, N., Beutels, P., Leirs, H. & Reijniers, J. Estimating Time of Infection Using Prior Serological and Individual Information Can Greatly Improve Incidence Estimation of Human and Wildlife Infections. PLOS Comput. Biol. 12, e1004882 (2016).

21. Greenhouse, B., Smith, D. L., Rodríguez-Barraquer, I., Mueller, I. & Drakeley, C. J. Taking Sharper Pictures of Malaria with CAMERAs: Combined Antibodies to Measure Exposure Recency Assays. Am. J. Trop. Med. Hyg. 99, 1120–1127 (2018).

22. Greenhouse, B. et al. Priority use cases for antibody-detecting assays of recent malaria exposure as tools to achieve and sustain malaria elimination. Gates Open Research 3, 131 (2019).

23. White, M. T. et al. Dynamics of the antibody response to Plasmodium falciparum infection in African children. J. Infect. Dis. 210, 1115–1122 (2014).

24. Yman, V. et al. Antibody responses to merozoite antigens after natural Plasmodium falciparum infection: Kinetics and longevity in absence of re-exposure. BMC Med. 17, (2019).

25. Kinyanjui, S. M., Conway, D. J., Lanar, D. E. & Marsh, K. IgG antibody responses to Plasmodium falciparum merozoite antigens in Kenyan children have a short half-life. Malar. J. 6, (2007).

26. Helb, D. A. et al. Novel serologic biomarkers provide accurate estimates of recent Plasmodium falciparum exposure for individuals and communities. Proc. Natl. Acad. Sci. 112, E4438–E4447 (2015).

27. Perraut, R. et al. Serological signatures of declining exposure following intensification of integrated malaria control in two rural Senegalese communities. PLoS One 12, e0179146 (2017).

28. King, C. L. et al. Biosignatures of Exposure/Transmission and Immunity. Am. J. Trop. Med. Hyg. 93, 16–27 (2015).

29. Proietti, C. et al. Immune Signature Against Plasmodium falciparum Antigens Predicts Clinical Immunity in Distinct Malaria Endemic Communities. Mol. Cell. Proteomics 19, 101–113 (2020).

30. Helb, D. Anti-malarial Antibody Responses & Applications for Assessing Malaria Exposure. (UC Berkeley, 2015).

31. Kobayashi, T. et al. Distinct Antibody Signatures Associated with Different Malaria Transmission Intensities in Zambia and Zimbabwe. mSphere 4, (2019).

32. van den Hoogen, L. L. et al. Selection of Antibody Responses Associated With Plasmodium falciparum Infections in the Context of Malaria Elimination. Front. Immunol. 11, 928 (2020).

33. Longley, R. J. et al. Development and validation of serological markers for detecting recent Plasmodium vivax infection. Nat. Med. 2020 265 26, 741–749 (2020).

34. van den Hoogen, L. L. et al. Antibody Responses to Antigenic Targets of Recent Exposure Are Associated With Low-Density Parasitemia in Controlled Human Plasmodium falciparum Infections. Front. Microbiol. 9, 3300 (2018).

35. Wu, L. et al. Comparison of diagnostics for the detection of asymptomatic Plasmodium falciparum infections to inform control and elimination strategies. Nature 528, S86– S93 (2015).

36. Sheehy, S. H. et al. ChAd63-MVA-vectored blood-stage malaria vaccines targeting MSP1 and AMA1: assessment of efficacy against mosquito bite challenge in humans. Mol. Ther. 20, 2355–68 (2012).

37. Walker, K. M. et al. Antibody and T-cell responses associated with experimental human malaria infection or vaccination show limited relationships. Immunology 145, 71–81 (2015).

38. Scholzen, A. & Sauerwein, R. W. Immune activation and induction of memory: lessons learned from controlled human malaria infection with Plasmodium falciparum. Parasitology 143, 224–35 (2016).

39. Homann, M. V. et al. Detection of Malaria Parasites After Treatment in Travelers: A 12-months Longitudinal Study and Statistical Modelling Analysis. EBioMedicine 25, 66–72 (2017).

40. Asghar, M. et al. Cellular aging dynamics after acute malaria infection: A 12-month longitudinal study. Aging Cell 17, e12702 (2018).

41. Sundling, C. et al. B cell profiling in malaria reveals expansion and remodeling of CD11c+ B cell subsets. JCI Insight 4, (2019).

42. Kamuyu, G. et al. KILchip v1.0: A Novel Plasmodium falciparum Merozoite Protein Microarray to Facilitate Malaria Vaccine Candidate Prioritization. Front. Immunol. 9, 2866 (2018).

43. World Health Organization. Disease surveillance for malaria elimination?: an operational manual. (World Health Organization, 2012).

44. Drewe, J. A., Hoinville, L. J., Cook, A. J. C., Floyd, T. & Stärk, K. D. C. Evaluation of animal and public health surveillance systems: a systematic review. Epidemiol. Infect. 140, 575–590 (2012).

45. Groseclose, S. L. & Buckeridge, D. L. Public Health Surveillance Systems: Recent Advances in Their Use and Evaluation. Annu. Rev. Public Health 38, 57–79 (2017).

46. Guidelines Working Group. Updated Guidelines for Evaluating Public Health Surveillance Systems. (2001).

47. Avdicova, M. et al. Data quality monitoring and surveillance system evaluation: A handbook of methods and applications. ECDC Technical Document 1–91 (2014). doi:10.2900/35329

48. McCallum, F. J. et al. Differing rates of antibody acquisition to merozoite antigens in malaria: implications for immunity and surveillance. J. Leukoc. Biol. 101, 913–925 (2016).

49. Burel, J. G. et al. Dichotomous miR expression and immune responses following primary blood-stage malaria. JCI insight 2, (2017).

50. Stewart, L. et al. Rapid assessment of malaria transmission using age-specific sero-conversion rates. PLoS One 4, e6083 (2009).

51. Weber, G. E. et al. Sero-catalytic and Antibody Acquisition Models to Estimate Differing Malaria Transmission Intensities in Western Kenya. Sci. Rep. 7, 16821 (2017).

52. Perraut, R. et al. Association of antibodies to Plasmodium falciparum merozoite surface protein-4 with protection against clinical malaria. Vaccine 35, 6720–6726 (2017).

53. Adu, B. et al. Antibody levels against GLURP R2, MSP1 block 2 hybrid and AS202.11 and the risk of malaria in children living in hyperendemic (Burkina Faso) and hypoendemic (Ghana) areas. Malar. J. 15, (2016).

54. Landier, J. et al. Operational performance of a plasmodium falciparum ultrasensitive rapid diagnostic test for detection of asymptomatic infections in eastern Myanmar. J. Clin. Microbiol. 56, (2018).

55. Mwesigwa, J. et al. Field performance of the malaria highly sensitive rapid diagnostic test in a setting of varying malaria transmission. Malar. J. 18, 288 (2019).

56. Longley, R. J. et al. Naturally acquired antibody responses to more than 300 Plasmodium vivax proteins in three geographic regions. PLoS Negl. Trop. Dis. 11, e0005888 (2017).

57. Sboner, A. et al. Robust-Linear-Model Normalization To Reduce Technical Variability in Functional Protein Microarrays research articles. J. Proteome Res. 8, 5451–5464 (2009).

58. Sill, M., Schröder, C., Hoheisel, J. D., Benner, A. & Zucknick, M. Assessment and optimisation of normalisation methods for dual-colour antibody microarrays. BMC Bioinformatics 11, 556 (2010).

59. Benjamini, Y. & Hochberg, Y. Controlling the False Discovery Rate: A Practical and Powerful Approach to Multiple Testing. Journal of the Royal Statistical Society. Series B (Methodological) 57, 289–300 (1995).

60. Sun, X. & Xu, W. Fast implementation of DeLong’s algorithm for comparing the areas under correlated receiver operating characteristic curves. IEEE Signal Process. Lett. 21, 1389–1393 (2014).

61. Nilsson, R., Peña, J. M., Björkegren, J. & Tegnér, J. Consistent Feature Selection for Pattern Recognition in Polynomial Time. J. Mach. Learn. Res. 8, 589–612 (2007).

62. Kursa, M. B., Rudnicki, W. R., Hastie, T., Tibshirani, R. & Friedman, J. Feature selection with the Boruta Package. J. Stat. Softw. 36, (2010).

63. Dubitzky, Werner; Granzow, Martin; Berrar, D. Fundamentals of data mining in genomics and proteomics. (Springer Science & Business Media., 2007).

64. White, M. et al. Antibody kinetics following vaccination with MenAfriVac: an analysis of serological data from randomised trials. Lancet Infect. Dis. 19, 327–336 (2019).

