## Supplementary material for "Distinct kinetics in antibody responses to 111 *Plasmodium falciparum* antigens identifies novel serological markers of recent malaria exposure": File S1

Supplementary Information for:

### Distinct kinetics in antibody responses to 111 *Plasmodium falciparum* antigens identifies novel serological markers of recent exposure

Yman V, Tuju J, White M T, Kamuyu G, Mwai K, Kibinge N, Asghar M, Sundling C, Sondén K, Murungi L, Kiboi D, Kimathi R, Chege T, Chepsat E, Kiyuka P, Nyamako L, Osier F H A, Färnert A

### Supplementary Tables

**Supplementary Table S1.** Descriptive statistics of the study participants.

| | Primary infected | Previously exposed |
|---|---|---|
| Number of participants | 21 | 44 |
| Female sex (%) | 4 (19) | 7 (23) |
| Median age, years (range) | 34 (21-59) | 40 (27-70) |
| Median cumulative time of residency in endemic area, years (range) | 0 (0-3) | 25 (13-39) |
| Median time since residency in endemic area, years (range) | - | 14 (0-46) |
| Median time from symptom onset to diagnosis, days (range) | 3 (0-11) | 3 (1-13) |
| Median parasitaemia, % infected RBCs (range) | 0.45 (<0.1-8.0) | 0.3 (<0.1-7.6) |
| Late treatment failure* (%) | 5 (25) | 0 (0) |
| Severe malaria† (%) | 1 (5) | 4 (9.7) |
| Treated in intensive care unit (%) | 1 (5) | 2 (4.9) |
| Initial intraveneous artesunate treatment (%) | 6 (30) | 10 (24.4) |

* Presented with recrudescent parasitaemia and fever 20-28 days following initial treatment.

† Severe malaria was defined according to the WHO criteria which include impaired consciousness, acidosis, hypoglycaemia, severe anaemia, renal impairment, jaundice, pulmonary oedema, bleeding, circulatory shock and hyperparasitemia (Management of Severe Malaria: A Practical Handbook, 3rd edition, 1-83, World Health Organization, 2012).

### Supplementary Figures

#### Supplementary Fig. S1.

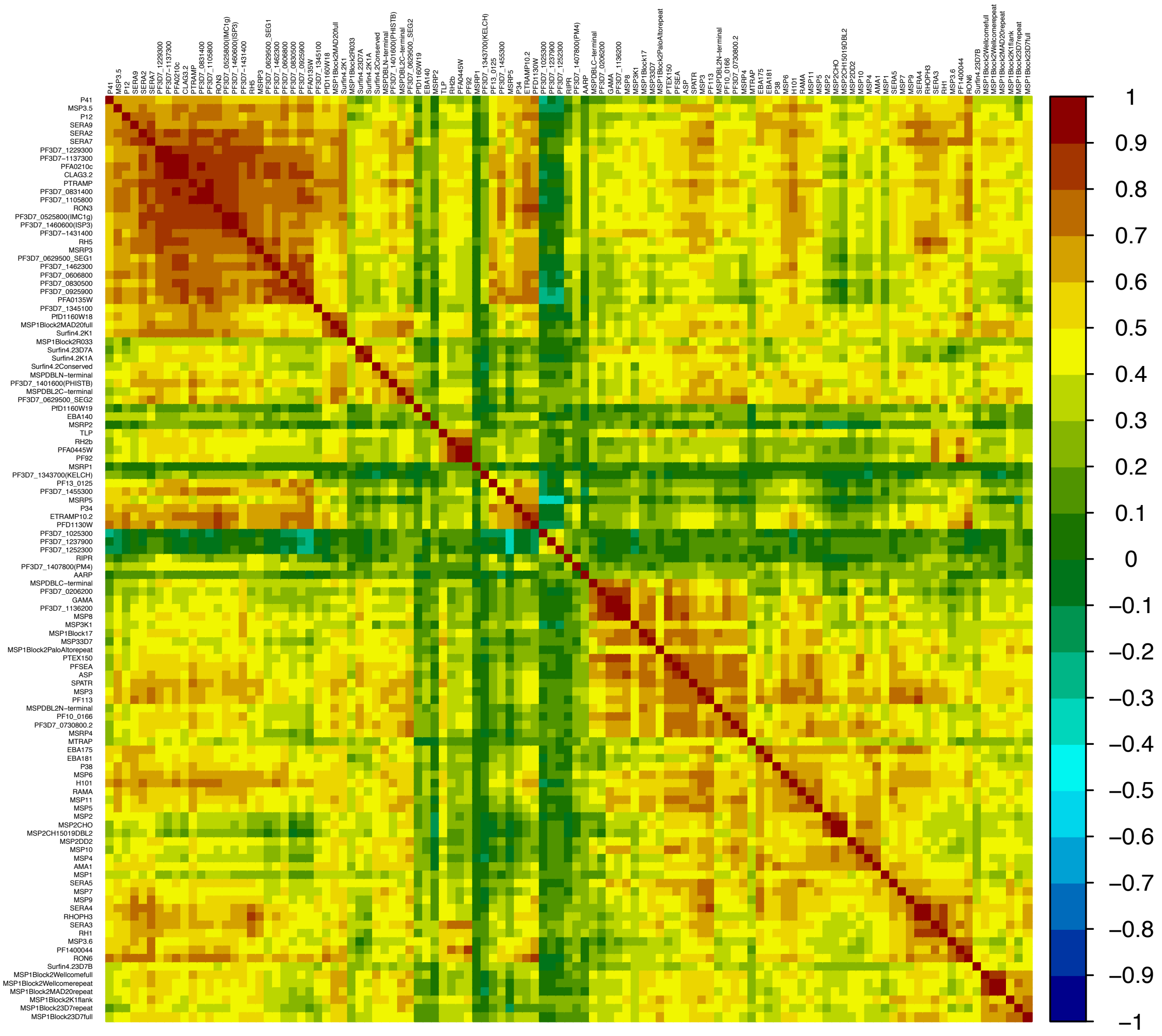


**Heatmap displaying the correlation of all pairwise comparisons of antibody responses (n = 6105).** Positive correlation is indicated in red while negative correlation is indicated in blue. The magnitude of the correlation is given by the colour intensity. Responses are ordered using a hierarchical clustering based on the magnitude of the correlation.

### Supplementary Fig. S2

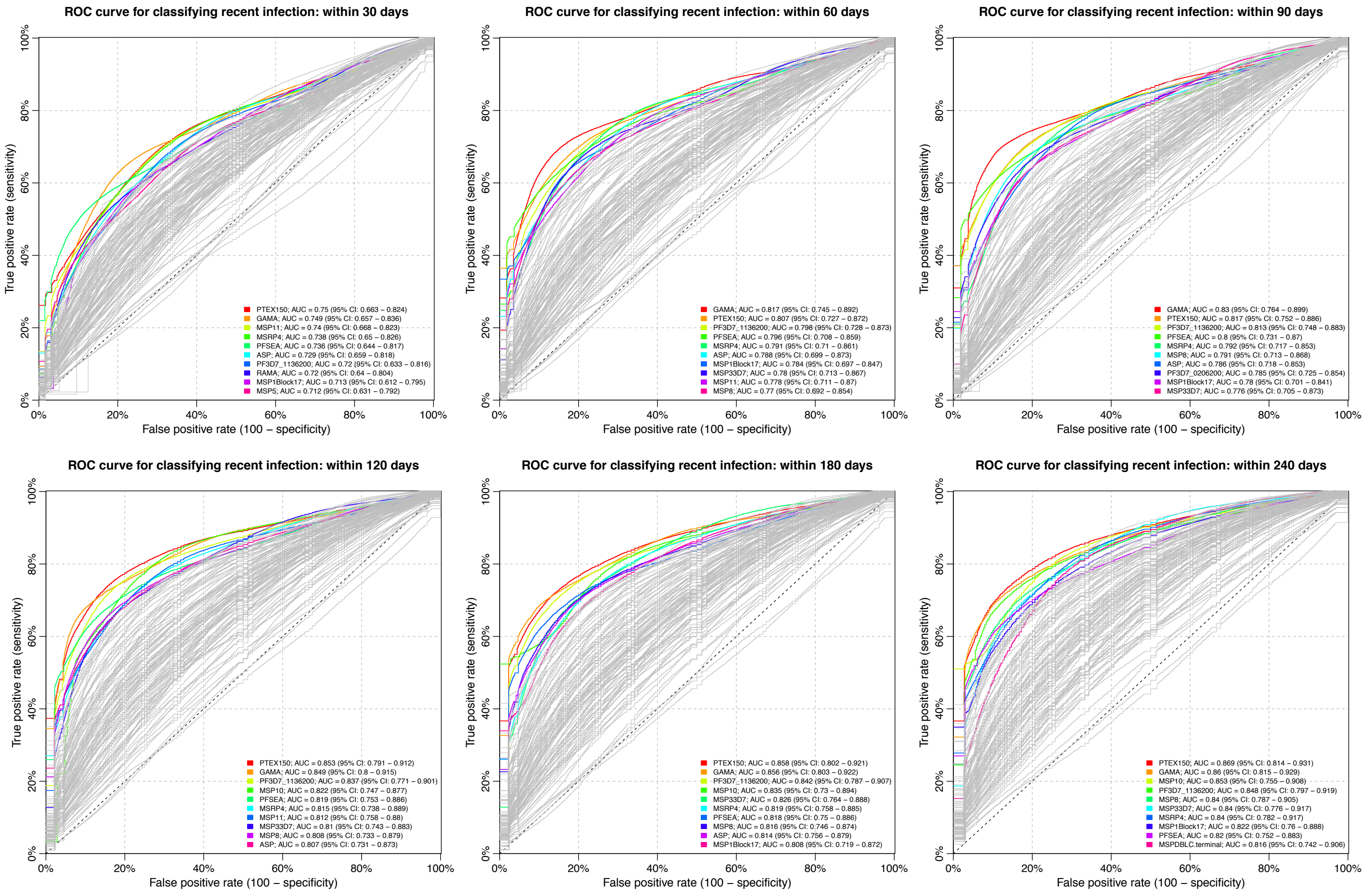


**Receiver operating characteristic (ROC) curve for classifying individuals as recently infected using a threshold antibody level to a single antigen.** The analysis was repeated for a range of temporal thresholds used to define a recent infection (i.e. 1, 2, 3, 4, 6, and 8 months) and each panel represents the results for a given temporal threshold. Coloured curves in each panel correspond to the top 10 antibody responses that were most predictive of recent infection as determined by the classifier area under the ROC curve (AUC).

### Supplementary Fig. S3

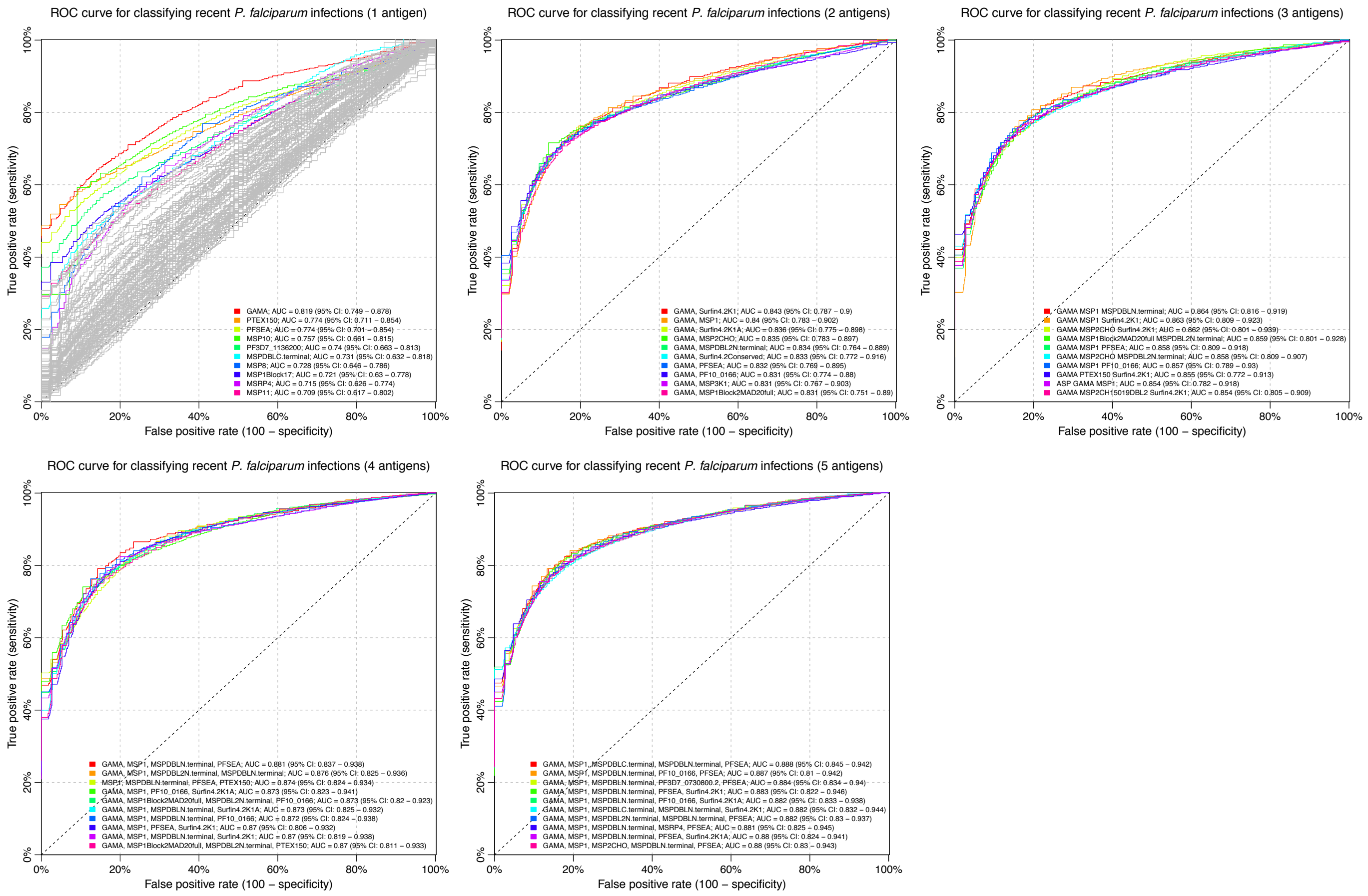


**Cross-validated ROC-curves of classifier performance for random classifiers fitted to data on antibody responses towards a combination of two to five out of the 28 selected antigens.** There was a gradual increase in random forest classifier performance with the inclusion of increasing number (1 to 5) of antibody responses. Individual panels represent the cross-validated receiver operating characteristic (ROC) curves for random forest classifiers fitted to data on antibody responses to the top 10 combinations of one (111), two (379 combinations), three (3276 combinations), four (20475 combinations), and five (98280 combinations) antibody responses out of the 28 selected, respectively. The classifier performance was evaluated using the area under the ROC curve (AUC).

### Supplementary Fig S4

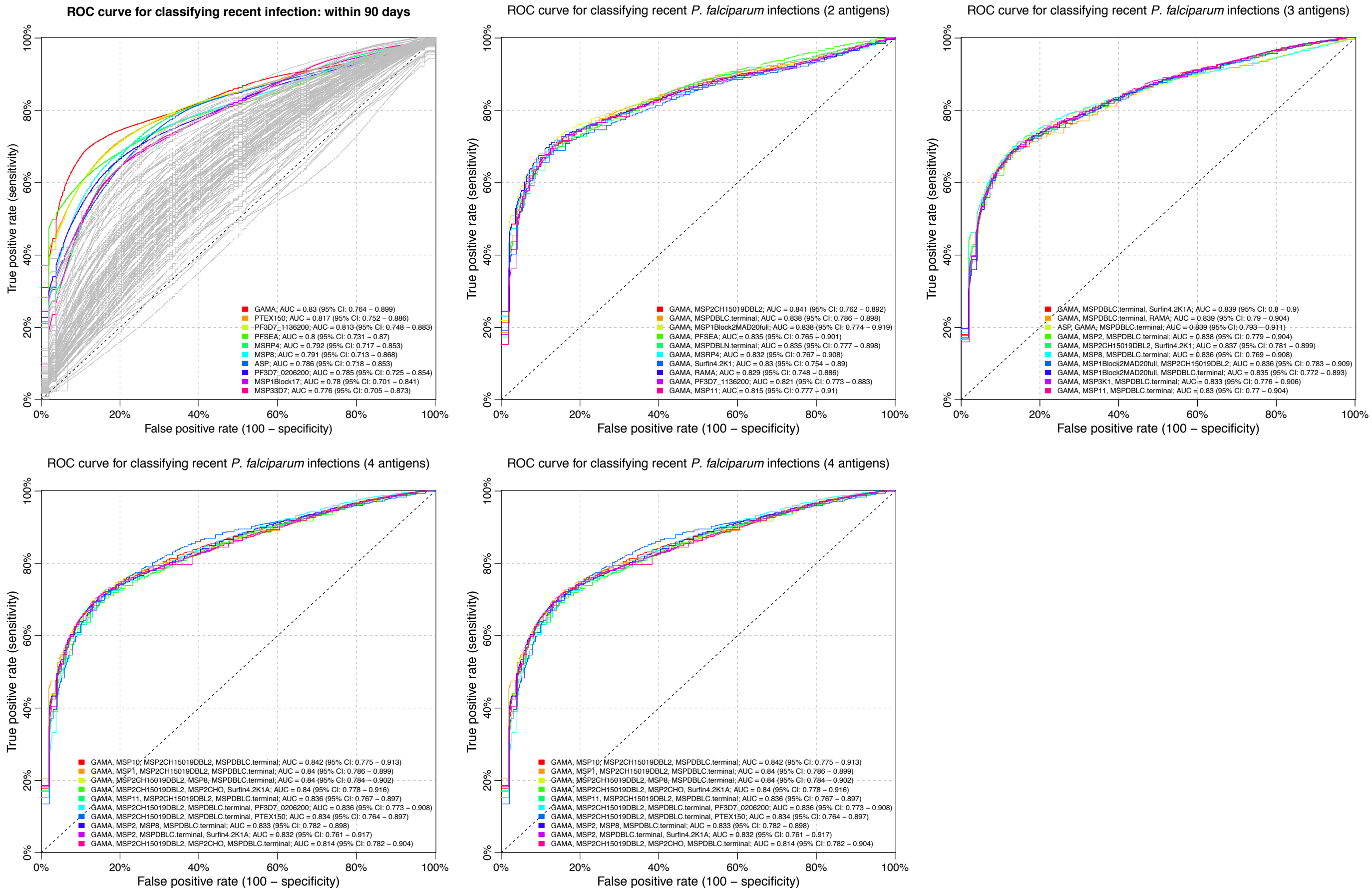


**Cross-validated ROC-curves of classifier performance for logistic regression classifiers fitted to data on antibody responses towards a combination of two to five out of the 28 selected antigens.** There was no significant increase in logistic regression classifier performance with the inclusion of increasing number (1 to 5) of antibody responses. Individual panels represent the cross-validated receiver operating characteristic (ROC) curves for logistic regression classifiers fitted to data on antibody responses to the top 10 combinations of one (111), two (379 combinations), three (3276 combinations), four (20475 combinations), and five (98280 combinations) antibody responses out of the 28 selected, respectively. The classifier performance was evaluated using the area under the ROC curve (AUC).

**Supplementary Fig S5**

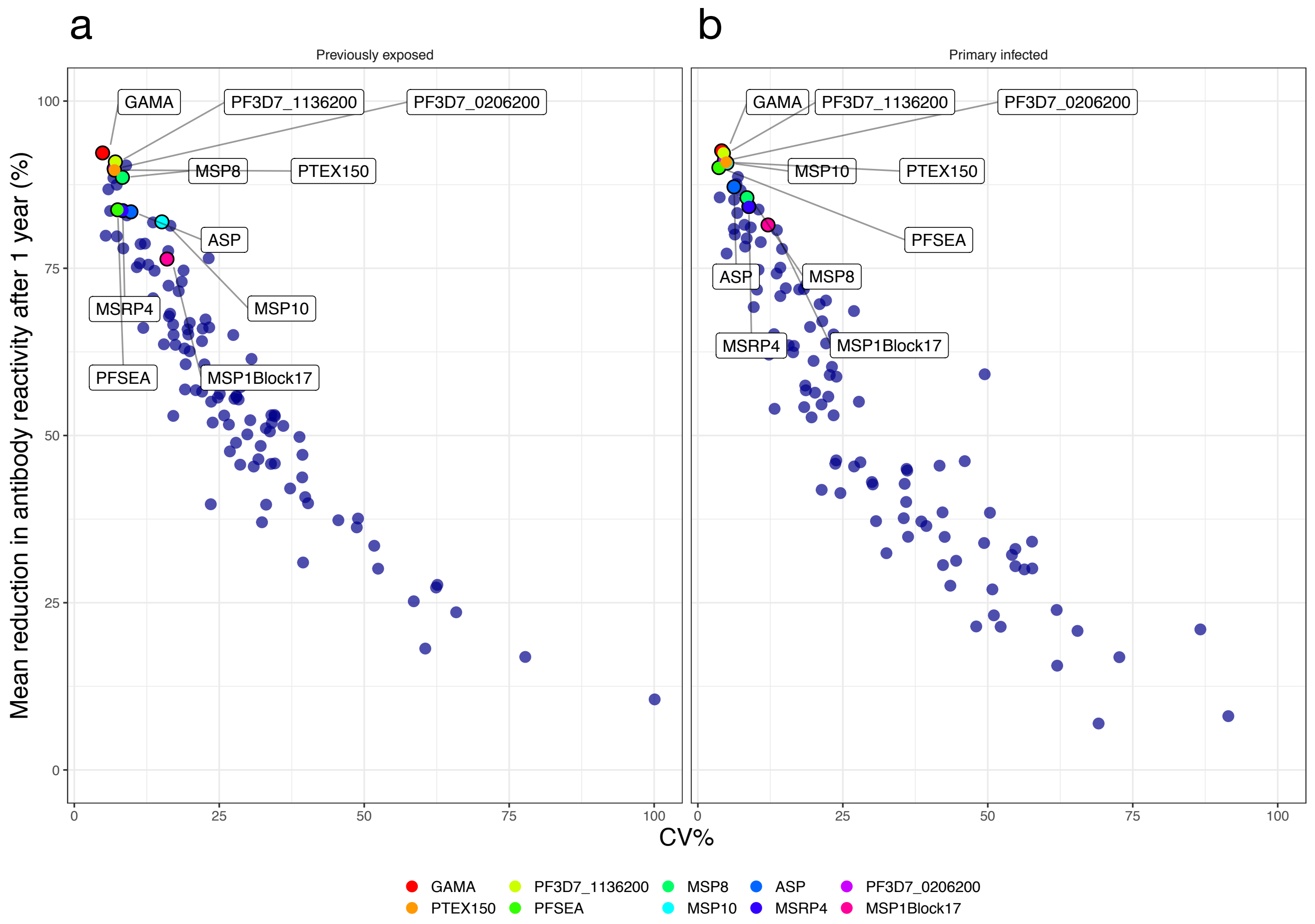


**Mean relative reduction in antibody levels after 1 year of follow-up versus the coefficient of variation of the estimated relative reduction in (a) previously exposed and (b) primary infected individuals, respectively.** Colours indicate the antibody responses identified as top 10 most informative in detecting recent infection based on a threshold antibody level to a single antigen.
